# The Effects of Psychosocial Stress on Memory and Cognitive Ability: A Meta-Analysis

**DOI:** 10.1101/2020.11.30.20240705

**Authors:** Elizabeth McManus, Deborah Talmi, Hamied Haroon, Nils Muhlert

**Author notes:** **Corresponding Author:** Dr Nils Muhlert, The University of Manchester, Zochonis building, Oxford Rd, Manchester, M13 9PL.

## Abstract

The impact of stress on cognitive abilities, such as memory, is well documented in animal studies but it is not yet clear how stress in human social interactions affects memory. This review systematically explored the evidence regarding the effects of psychosocial stress on memory and associated cognitive abilities. PubMed, PsycInfo and Web of Science databases were searched for studies assessing the effects of psychosocial stress on long-term memory or related cognitive functions. Fifty-one studies were identified and compared based on the timing of stress induction. No overall effect of psychosocial stress induction was seen on long-term or working memory regardless of whether stress induction occurred following encoding or before retrieval. Psychosocial stress had a moderate effect in studies comparing memory for emotional compared to neutral stimuli, but the direction of this effect varied across studies. Psychosocial stress decreased performance on executive function tasks. Our findings demonstrate that psychosocial stress may not have the clear effects on memory previously ascribed to it, suggesting potentially different mechanisms from physiological stressors.

## 1. Introduction

Many people report memory failures when trying to retrieve previously learnt information in times of high stress, such as when giving important presentations or taking exams. In contrast, memory for events that occurred in a moderately emotive or stressful environment are often reported to be enhanced. This dichotomous relationship between the influences of stress on cognitive processes is becoming increasingly important given the prevalence of high levels of social stress experienced in our daily lives (Ravalier & Walsh, 2018) and the necessity for intact cognitive ability for a good quality of physical, mental and social well-being (Bailey, 2007; Brown & Landgraf, 2010; Diamond, 2013). Reviews of studies in human have often focussed mainly on physiological stressors (such as cold water challenges), but evidence suggests that the neural pathways involved in psychosocial and physiological stress differ (Dedovic, Duchesne, Andrews, Engert, & Pruessner, 2009). In addition, the impact of psychosocial stress is influenced by the social context of an individual, further suggesting differences in the effects of these forms of stress. Given that these experimental studies are typically interpreted in the light of stresses in everyday life it therefore makes sense to understand how, if at all, psychosocial stress affects cognition.

### 1.1 Neurobiological underpinnings of the effects of stress on memory

Acute stress, through associated elevations in stress-related hormones, can impact cognitive abilities, including memory, attention and cognitive flexibility (McEwen & Sapolsky, 1995; Olver, Pinney, Maruff, & Norman, 2015; Roozendaal, McEwen, & Chattarji, 2009). However, the timing of stress has been argued to impact on how stress affects cognition (Shields, Sazma, McCullough, & Yonelinas, 2017; Tom Smeets, Otgaar, Candel, & Wolf, 2008; Wolf, 2009). Stress during encoding has been shown to facilitate learning and enhance subsequent memory retrieval (Buchanan & Lovallo, 2001; Cahill, Gorski, & Le, 2003; Joëls, Pu, Wiegert, Oitzl, & Krugers, 2006). In contrast, stress immediately before retrieval is reported to impair memory (Smeets, 2011; Wolf, 2017).

The relationship between stress and memory is thought to be mediated by glucocorticoids and catecholamines, that are implicated in both stress-related memory enhancement and impairment (Het, Ramlow, & Wolf, 2005; Schwabe, Joëls, Roozendaal, Wolf, & Oitzl, 2012). The proposed neurobiological basis for the influence of stress on memory suggests overlap between neural circuits engaged in responding to stress-related hormones and neural circuits involved in memory (Joëls et al., 2006). Experimental evidence in animals has demonstrated that glucocorticoids enhance memory consolidation through their interactions with the basolateral amygdala, impacting on memory processes in the hippocampus (Roozendaal et al., 2009; Roozendaal, Nguyen, Power, & McGaugh, 1999; Roozendaal, Williams, & McGaugh, 1999). In contrast to this enhancing effect, when present at retrieval, glucocorticoids are suggested to impair hippocampal-dependent memory retrieval (Gagnon & Wagner, 2016; Hargis, Buechel, Popovic, & Blalock, 2018; Roozendaal, 2002; Wolf, 2017; Zoladz, Park, Fleshner, & Diamond, 2015). These animal studies suggest that stressors influence hippocampal function, indicating a shared biological mechanism for the effects of stress on memory (Fuchs et al., 2001; Ohl, Michaelis, Vollmann-Honsdorf, Kirschbaum, & Fuchs, 2000). In humans however, there is evidence that different forms of stress, such as physical, cognitive or psychosocial stressors may differ in their effects on specific neurobiological pathways (Alleva & Santucci, 2001; Dedovic et al., 2009; Joëls et al., 2006). During physical pain, human brain activity increases in the insula, thalamus and motor-related regions of the brain. In contrast, psychosocial stressors elicit activation in the limbic system, including the hippocampus and amygdala (Herman & Cullinan, 1997; Peyron, Laurent, & Garcia-Larrea, 2000). These differences in regional activity may result in stressor-specific cognitive changes. In particular, psychosocial stress may particularly impact upon episodic memory processes that are known to rely on the hippocampus (McGaugh, 2000, 2004; Yonelinas & Ritchey, 2015).

### 1.2 Retrieval of emotional and neutral memories

Intrinsic features of stimuli, such as emotional valence, may also help to determine the likelihood of retrieval. Memory for emotional stimuli is typically better than memory for neutral stimuli (LaBar & Cabeza, 2006; Ponzio & Mather, 2014; Sharot & Yonelinas, 2008; Talmi, 2013). This phenomenon can be explained using the modulation model: emotional memories are less likely to be forgotten than neutral memories as they are associated with greater engagement of the amygdala at encoding (McGaugh, 2004). Secretion of noradrenergic and other stress-related hormones, such as cortisol, due to the arousal evoked by emotional stimuli may further enhance this amygdala activation at encoding (McGaugh, 2004).

Building on this model, mediation theory suggests that emotional stimuli will be better retrieved than neutral stimuli before consolidation has occurred (Talmi, 2013). This model posits that the emotional memory advantage results from the increased distinctiveness of emotional stimuli (Schmidt & Saari, 2007; Schmidt & Schmidt, 2016) and by its inherent ability to attract more attention than neutral stimuli (Talmi & McGarry, 2012; Talmi, Schimmack, Paterson, & Moscovitch, 2007). in addition, the superior organisation and formation of greater semantic links for emotional compared to neutral stimuli offer further advantages for retention and retrieval (Talmi & Moscovitch, 2004).

### 1.3 Effects of stress on other cognitive functions

Stress is thought to impair executive functions, such as working memory and cognitive flexibility (Diamond, 2013; Shields, Sazma, & Yonelinas, 2016). Stress-related effects on memory may therefore be influenced, at least in part, by effects on other forms of cognition. The most prevalent theory suggests that stress may bias cognitive resources towards dealing with the current stressor, thus limiting available resources for other cognitive processes (LeBlanc, 2009; Sänger, Bechtold, Schoofs, Blaszkewicz, & Wascher, 2014; Schoofs, Wolf, & Smeets, 2009).

Selective and sustained attention is critical for effective memory retrieval, in order for individuals to recall specific information (Dudukovic, DuBrow, & Wagner, 2009). These aspects of attention may however be negatively affected by stress (Sänger et al., 2014). Additionally, working memory, a temporary storage that holds information for processing, has been demonstrated to be negatively influenced by stress (Oei, Everaerd, Elzinga, Van Well, & Bermond, 2006) on verbal and digit span and n-back tasks (Olver et al., 2015; Schoofs, Preuß, & Wolf, 2008a; Schoofs et al., 2009).

### 1.4 Aims

Previous reviews (Shields et al., 2017; Wolf, 2009) highlight the differential effects of stress on different stages of memory, but these conflate the different forms of stressor. As discussed, there is reason to suggest that psychosocial stress differs from physical stress, both in its neurobiological underpinnings and how it interacts with other inter-individual differences. No prior memory review has looked at psychosocial stress in isolation. Here, we focus specifically on studies using psychosocial stressors in healthy adults to examine the following questions: first, does acute psychosocial stress differentially impact learning and retrieval; second, does psychosocial stress impact emotional memory to a greater extent than neutral stimuli; and third, does acute psychosocial stress affect other cognitive abilities, such as executive function and working memory.

To answer the first question, we compared peer-reviewed studies grouped by the time of stress induction. We considered whether psychosocial stress studies took place in a single session, such that the stressor could affect both encoding and retrieval (Smeets, Jelicic, & Merckelbach, 2006; Wolf, Schommer, Hellhammer, McEwen, & Kirschbaum, 2001), or studies where encoding and retrieval were assessed on separate days to allow for clearer separation of the effects of stress on these stages of memory (Cornelisse, van Stegeren, & Joëls, 2011; Smeets, Giesbrecht, Jelicic, & Merckelbach, 2007). To explore the second aim, we compared studies using both emotional and neutral stimuli. Finally, we assessed studies exploring the impact of psychosocial stress on other cognitive abilities, such as working memory and executive functions, which may impact on long-term memory.

## 2. Method

### 2.1 Search strategy

This review was conducted in accordance with the PRISMA guidelines (Liberati et al., 2009). The search was conducted on PubMed, PsycINFO and Web of Science and last updated in November 2020. This review examines studies of this topic, up to this date. After initial searches and extraction was completed, we also searched the reference lists of included papers for any other relevant material that may have been missed.

Search terms used were informed by common terminologies used throughout the wider stress and memory literature and additional filters for human adult studies and not animal or child studies were included. The terms used were: (Acute stress OR Psychological stress OR Social stress OR Trier social stress test OR Stress reactivity) AND (memory OR free recall OR recognition OR recollection) AND (humans AND adults).

### 2.2 Eligibility Criteria

Our inclusion criteria for considering studies were that: First, studies had to be published in a peer reviewed journal. Second, studies had to be carried out in young and middle aged adult (18-40yrs) human participants without neurological or psychiatric conditions, to minimise the effects of age-related or pathological change in cognitive ability. Third, papers must have been written in English. Fourth, studies had to include an acute psychosocial stress test condition and a separate control condition or in control group. Fifth, studies had to examine the effects of psychosocial stress on either episodic memory or other cognitive ability (i.e. executive functions or working memory). Finally, we only included studies carried out in controlled lab environments.

### 2.3 Exclusion Criteria

The exclusion criteria also set boundaries for publication, population, intervention and comparators. To meet these criteria, the following studies were excluded: studied published as a conference abstract, presentation or poster; studies carried out in animals; studies examining the effects of physical stress, such as pain or exercise (e.g. the cold pressor task); studies using drug administration to induced cortisol elevations; studies which did not compare the effects of psychosocial stress between experimental and control groups or conditions; studies using stress outside a controlled environment. Additionally, studies that did not provide enough information to compute an effect size for a comparison between a single timing of psychosocial stress and a control group were excluded if the data was still unavailable after attempts were made to contact the author.

### 2.4 Data Extraction

The results of the searches were first screened by title and abstract and then by full text. An independent reviewer also screened a subset of full texts to assess reviewer reliability. The percentage agreement between reviewers was 100%.

During full text screenings, any papers excluded were reported in an exclusion table along with the reason for their exclusion. Key information from all studies included was reported in a summary table. This information included 1) Author, year & country 2) Sample size 3) Stress test used 4) Memory testing used 5) cognitive function test used 6) key findings.

### 2.5 Quality Assessment

Quality assessment was carried out using an adapted version of the Critical appraisal skills programme (CASP) checklist to asses potential sources of bias in study designs, participant selection, and procedure and data analysis (Singh, 2013). The study design used in all the extracted papers was a cohort design; therefore, quality was assessed based on the main criteria points including: representativeness of the experimental group, identification of confounds and minimisation of bias. Quality assessment was used to ensure that findings and patterns emerging in the data are neither inflated nor hindered by poorer-quality studies.

### 2.6 Stress and memory phases: definitions

We examined the effects of stress on either encoding, maintenance or retrieval (or a combination of these stages) by classifying the timing of stress in the following ways:

i. *Pre-learning stress with a short retention interval* Studies using a single testing session, lasting between 60-90minutes, in which participants underwent a stress test, before encoding and retrieval tasks.
ii. *Post learning stress with a short retention interval* Studies using a single testing session lasting between 60-90minutes, in which participants encoded stimuli before undergoing a stress test, with the retrieval task occurring immediately after the stress test.
iii. *Pre-learning stress with a long retention interval* Studies with a stress test followed by encoding task, with at least a 24-hour delay before the retrieval task.
iv. *Post-encoding stress with a long retention interval* Studies examining the effects of a stress task that was administered shortly after the encoding task with at least a 24-hour delay period before the retrieval task.
v. *Stress immediately before retrieval* Studies using two testing sessions, in which the first has no stressor and either encoding tasks only, or encoding and an immediate retrieval task. The second session then involves a stress test followed by a delayed retrieval task. Three studies used a 24hour delay period, a fourth used a 135minute delay period. We interpret this latter study with caution due to the shorter retention period.
vi. *Stress during encoding* Studies that tested participants on information that was learnt while undergoing stress.

### 2.7 Meta-Analysis & Computation of Effect Sizes

The effect size measure we used was Cohen’s d standardised mean difference between the stress condition and the control condition. We decided not to conduct a meta-analysis on any grouping with less than five studies, because comparisons with fewer studies may be misleading (Valentine, Pigott, & Rothstein, 2010). To address our first aim, we divided studies based on the timing of stress induction. Three studies had examined post-encoding stress with long retention intervals (category iv) and only two studies had examined stress during encoding (category vi), therefore these categories were excluded from this analysis. To address our second aim, we analysed all studies examining the effects of psychosocial stress on memory for emotional and neutral stimuli. Although it would be interesting to examine the interaction of the timing of stress (first aim) and emotionality of stimuli (second aim), there were too few studies in each of the resulting groupings to permit this analysis. To answer our third question, we examined how stress immediately impacts upon executive function and working memory. In total, we conducted seven meta-analysis: four exploring the effects of stress on individual memory phases (i.e. categories i-iii and v above), one exploring the effects on emotional stimuli, and two exploring the impact of stress on other cognitive abilities (executive function and working memory).

Wherever possible, Cohen’s d was calculated using means, standard deviations and effect sizes. However, when relevant information was not reported, effect sizes were calculated from F statistics or converted from other formats such as partial eta squared. If effect sizes could still not be calculated, corresponding authors were contacted to obtain the necessary information.

### 2.8 Heterogeneity analysis

An I^2^ statistic was calculated to estimate the proportion of variance explained by heterogeneity and inconsistency between studies within each proposed analysis (Higgins & Thompson, 2002). This was first calculated for all studies exploring the effect of psychosocial stress on memory, before splitting these studies based on the timing of stress. Any score above 75% is thought to indicate high heterogeneity of studies. Possible heterogeneous outliers were identified via visual inspection of forest plots and where appropriate were removed using post-hoc sensitivity analysis.

## 3. Results

A flowchart of each stage of the searches is shown below in figure 1, beginning with 5082 studies initially identified, and 51 included in the review.

**Figure 1.**
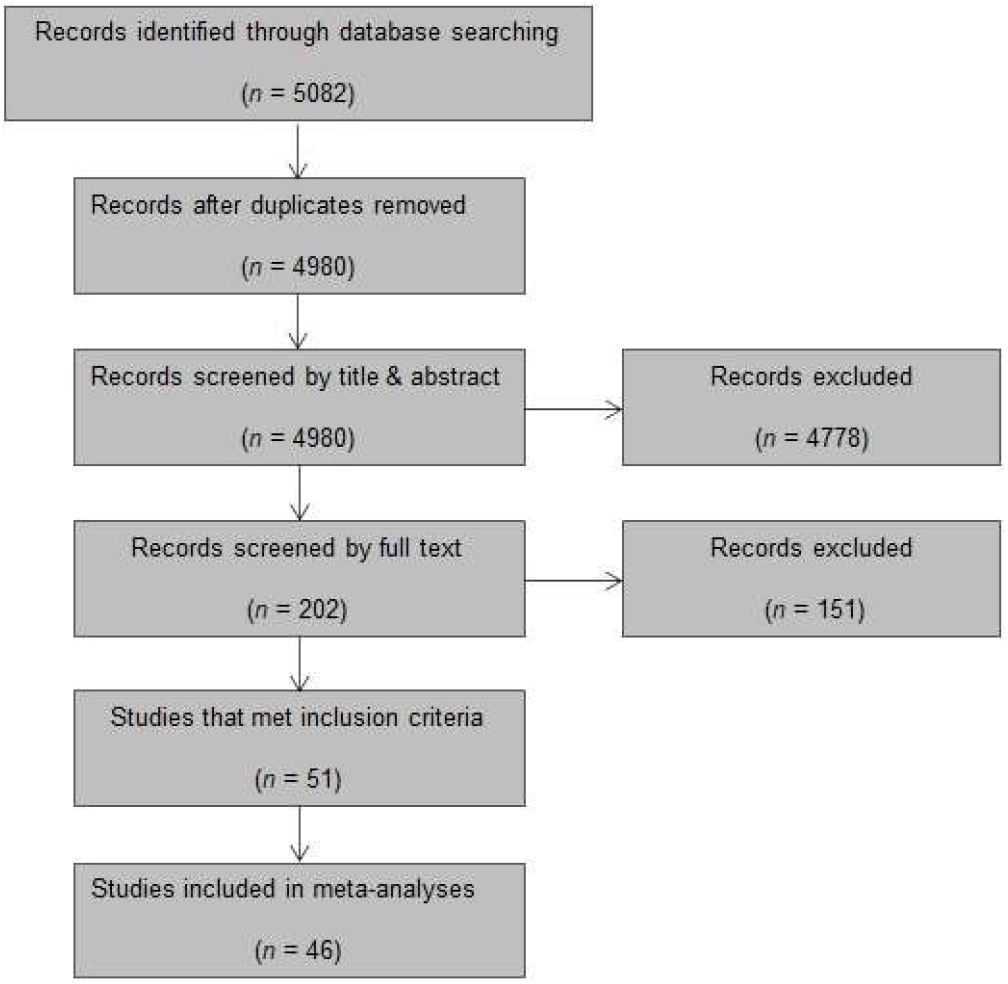
Studies Identified. PRISMA flow chart showing how many papers were selected or excluded at each stage.

### 3.1 Study Characteristics

A total of 51 papers were included in this review. Some papers included either multiple separate studies or have split analyses into males and females, so may be included multiple times in a meta-analysis. As this data is independent, no data correction was needed for these studies. Of the 51 papers included in the review, 29 assessed the effects of acute psychosocial stress on episodic memory, 17 on other cognitive functions, and 5 on both memory and other cognitive functions. Table 1 below summarises the characteristics and tasks used in the included studies assessing memory. Table 2 summarises the same features but for studies using other cognitive functions.

**Table 1.**
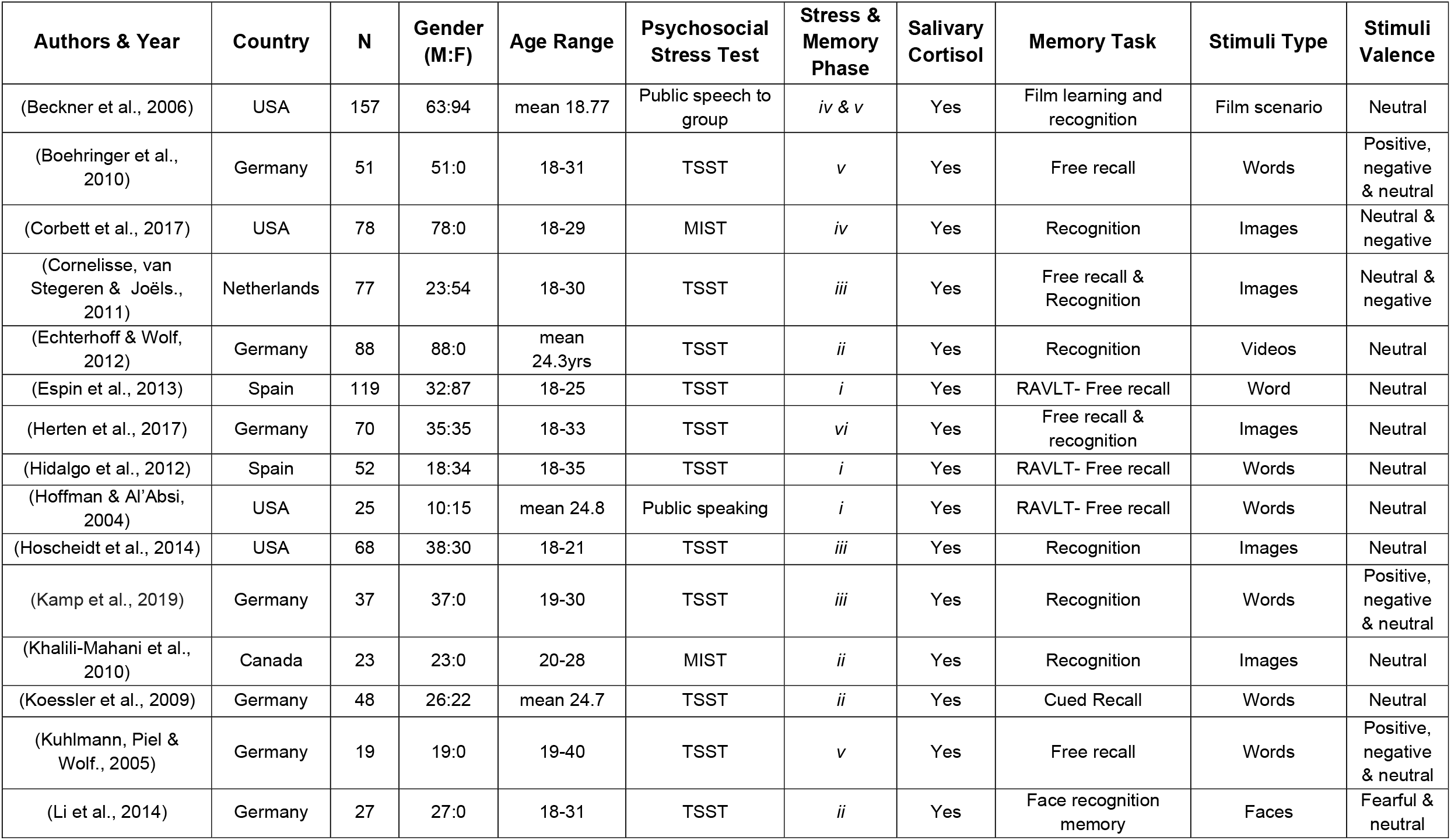
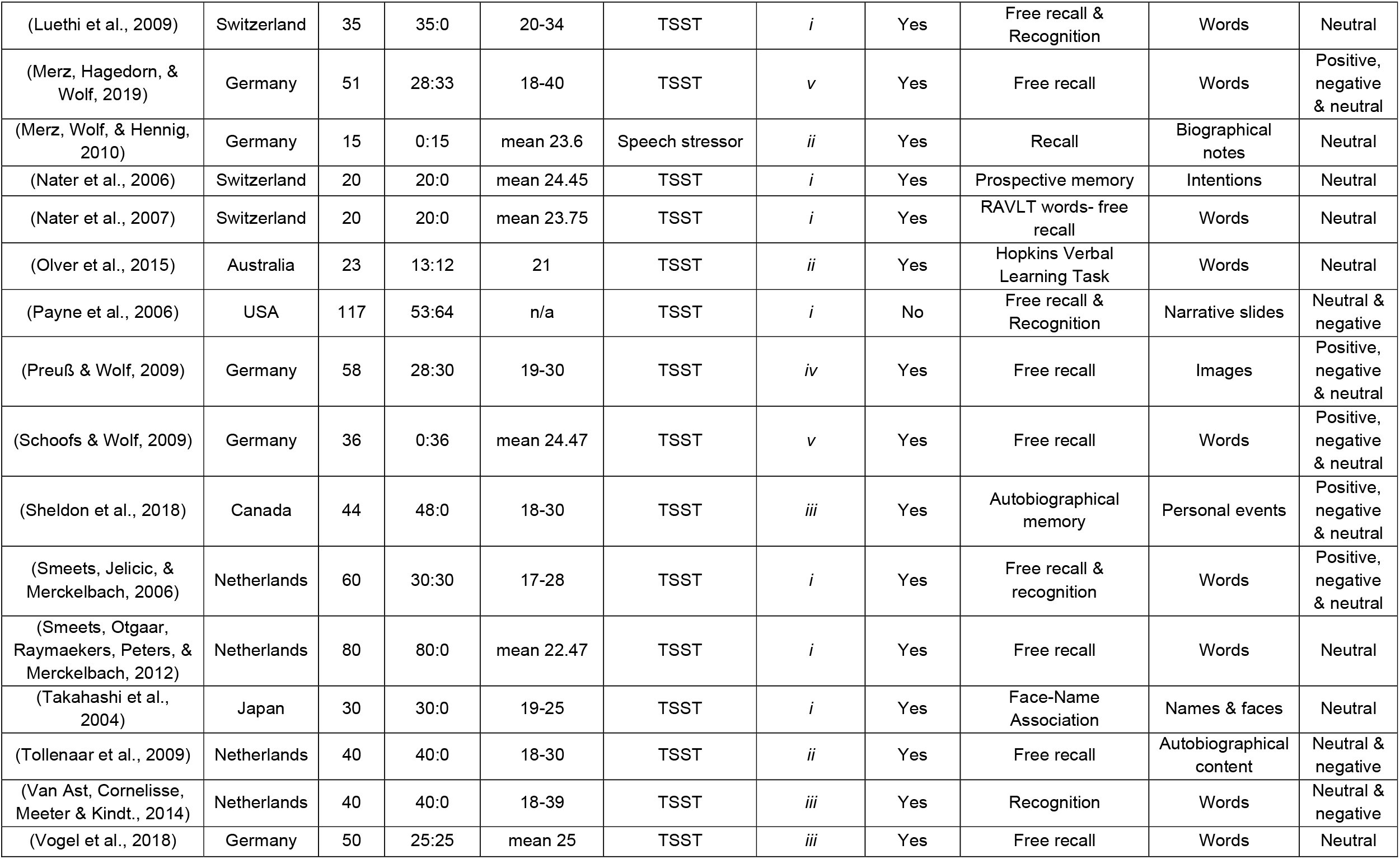
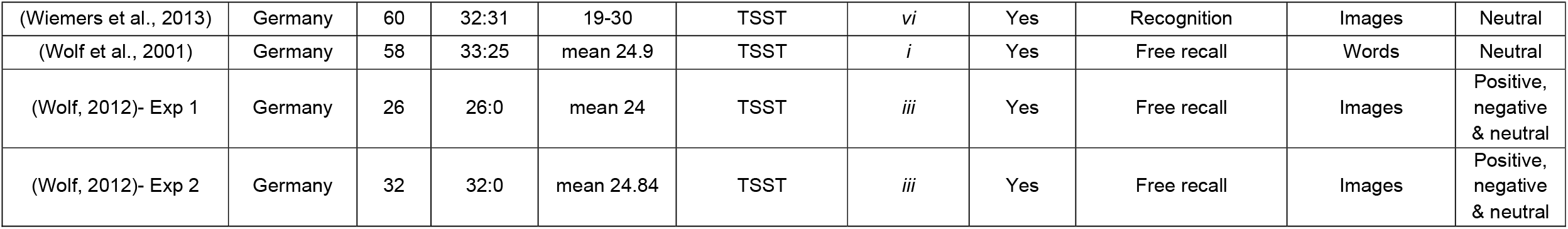
Study characteristics for all included studies using a memory task. Stress and memory phase column to the groupings defined above.

| Authors & Year | Country | N | Gender (M:F) | Age Range | Psychosocial Stress Test | Stress & Memory Phase | Salivary Cortisol | Memory Task | Stimuli Type | Stimuli Valence |
| --- | --- | --- | --- | --- | --- | --- | --- | --- | --- | --- |
| (Beckner et al., 2006) | USA | 157 | 63:94 | mean 18.77 | Public speech to group | *iv & v* | Yes | Film learning and recognition | Film scenario | Neutral |
| (Boehringer et al., 2010) | Germany | 51 | 51:0 | 18-31 | TSST | *v* | Yes | Free recall | Words | Positive, negative & neutral |
| (Corbett et al., 2017) | USA | 78 | 78:0 | 18-29 | MIST | *iv* | Yes | Recognition | Images | Neutral & negative |
| (Cornelisse, van Stegeren & Joëls., 2011) | Netherlands | 77 | 23:54 | 18-30 | TSST | *iii* | Yes | Free recall & Recognition | Images | Neutral & negative |
| (Echterhoff & Wolf, 2012) | Germany | 88 | 88:0 | mean 24.3yrs | TSST | *ii* | Yes | Recognition | Videos | Neutral |
| (Espin et al., 2013) | Spain | 119 | 32:87 | 18-25 | TSST | *i* | Yes | RAVLT- Free recall | Word | Neutral |
| (Herten et al., 2017) | Germany | 70 | 35:35 | 18-33 | TSST | *vi* | Yes | Free recall & recognition | Images | Neutral |
| (Hidalgo et al., 2012) | Spain | 52 | 18:34 | 18-35 | TSST | *i* | Yes | RAVLT- Free recall | Words | Neutral |
| (Hoffman & Al'Absi, 2004) | USA | 25 | 10:15 | mean 24.8 | Public speaking | *i* | Yes | RAVLT- Free recall | Words | Neutral |
| (Hoscheidt et al., 2014) | USA | 68 | 38:30 | 18-21 | TSST | *iii* | Yes | Recognition | Images | Neutral |
| (Kamp et al., 2019) | Germany | 37 | 37:0 | 19-30 | TSST | *iii* | Yes | Recognition | Words | Positive, negative & neutral |
| (Khalili-Mahani et al., 2010) | Canada | 23 | 23:0 | 20-28 | MIST | *ii* | Yes | Recognition | Images | Neutral |
| (Koessler et al., 2009) | Germany | 48 | 26:22 | mean 24.7 | TSST | *ii* | Yes | Cued Recall | Words | Neutral |
| (Kuhlmann, Piel & Wolf., 2005) | Germany | 19 | 19:0 | 19-40 | TSST | *v* | Yes | Free recall | Words | Positive, negative & neutral |
| (Li et al., 2014) | Germany | 27 | 27:0 | 18-31 | TSST | *ii* | Yes | Face recognition memory | Faces | Fearful & neutral |
| (Luethi et al., 2009) | Switzerland | 35 | 35:0 | 20-34 | TSST | *i* | Yes | Free recall & Recognition | Words | Neutral |
| (Merz, Hagedorn, & Wolf, 2019) | Germany | 51 | 28:33 | 18-40 | TSST | *v* | Yes | Free recall | Words | Positive, negative & neutral |
| (Merz, Wolf, & Hennig, 2010) | Germany | 15 | 0:15 | mean 23.6 | Speech stressor | *ii* | Yes | Recall | Biographical notes | Neutral |
| (Nater et al., 2006) | Switzerland | 20 | 20:0 | mean 24.45 | TSST | *i* | Yes | Prospective memory | Intentions | Neutral |
| (Nater et al., 2007) | Switzerland | 20 | 20:0 | mean 23.75 | TSST | *i* | Yes | RAVLT words- free recall | Words | Neutral |
| (Olver et al., 2015) | Australia | 23 | 13:12 | 21 | TSST | *ii* | Yes | Hopkins Verbal Learning Task | Words | Neutral |
| (Payne et al., 2006) | USA | 117 | 53:64 | n/a | TSST | *i* | No | Free recall & Recognition | Narrative slides | Neutral & negative |
| (Preuß & Wolf, 2009) | Germany | 58 | 28:30 | 19-30 | TSST | *iv* | Yes | Free recall | Images | Positive, negative & neutral |
| (Schoofs & Wolf, 2009) | Germany | 36 | 0:36 | mean 24.47 | TSST | *v* | Yes | Free recall | Words | Positive, negative & neutral |
| (Sheldon et al., 2018) | Canada | 44 | 48:0 | 18-30 | TSST | *iii* | Yes | Autobiographical memory | Personal events | Positive, negative & neutral |
| (Smeets, Jelicic, & Merckelbach, 2006) | Netherlands | 60 | 30:30 | 17-28 | TSST | *i* | Yes | Free recall & recognition | Words | Positive, negative & neutral |
| (Smeets, Otgaar, Raymaekers, Peters, & Merckelbach, 2012) | Netherlands | 80 | 80:0 | mean 22.47 | TSST | *i* | Yes | Free recall | Words | Neutral |
| (Takahashi et al., 2004) | Japan | 30 | 30:0 | 19-25 | TSST | *i* | Yes | Face-Name Association | Names & faces | Neutral |
| (Tollenaar et al., 2009) | Netherlands | 40 | 40:0 | 18-30 | TSST | *ii* | Yes | Free recall | Autobiographical content | Neutral & negative |
| (Van Ast, Cornelisse, Meeter & Kindt., 2014) | Netherlands | 40 | 40:0 | 18-39 | TSST | *iii* | Yes | Recognition | Words | Neutral & negative |
| (Vogel et al., 2018) | Germany | 50 | 25:25 | mean 25 | TSST | *iii* | Yes | Free recall | Words | Neutral |

|  |  |  |  |  |  |  |  |  |  |  |
| --- | --- | --- | --- | --- | --- | --- | --- | --- | --- | --- |
| (Wiemers et al., 2013) | Germany | 60 | 32:31 | 19-30 | TSST | *vi* | Yes | Recognition | Images | Neutral |
| (Wolf et al., 2001) | Germany | 58 | 33:25 | mean 24.9 | TSST | *i* | Yes | Free recall | Words | Neutral |
| (Wolf, 2012)- Exp 1 | Germany | 26 | 26:0 | mean 24 | TSST | *iii* | Yes | Free recall | Images | Positive,<br>negative<br>& neutral |
| (Wolf, 2012)- Exp 2 | Germany | 32 | 32:0 | mean 24.84 | TSST | *iii* | Yes | Free recall | Images | Positive,<br>negative<br>& neutral |

**Table 2.**
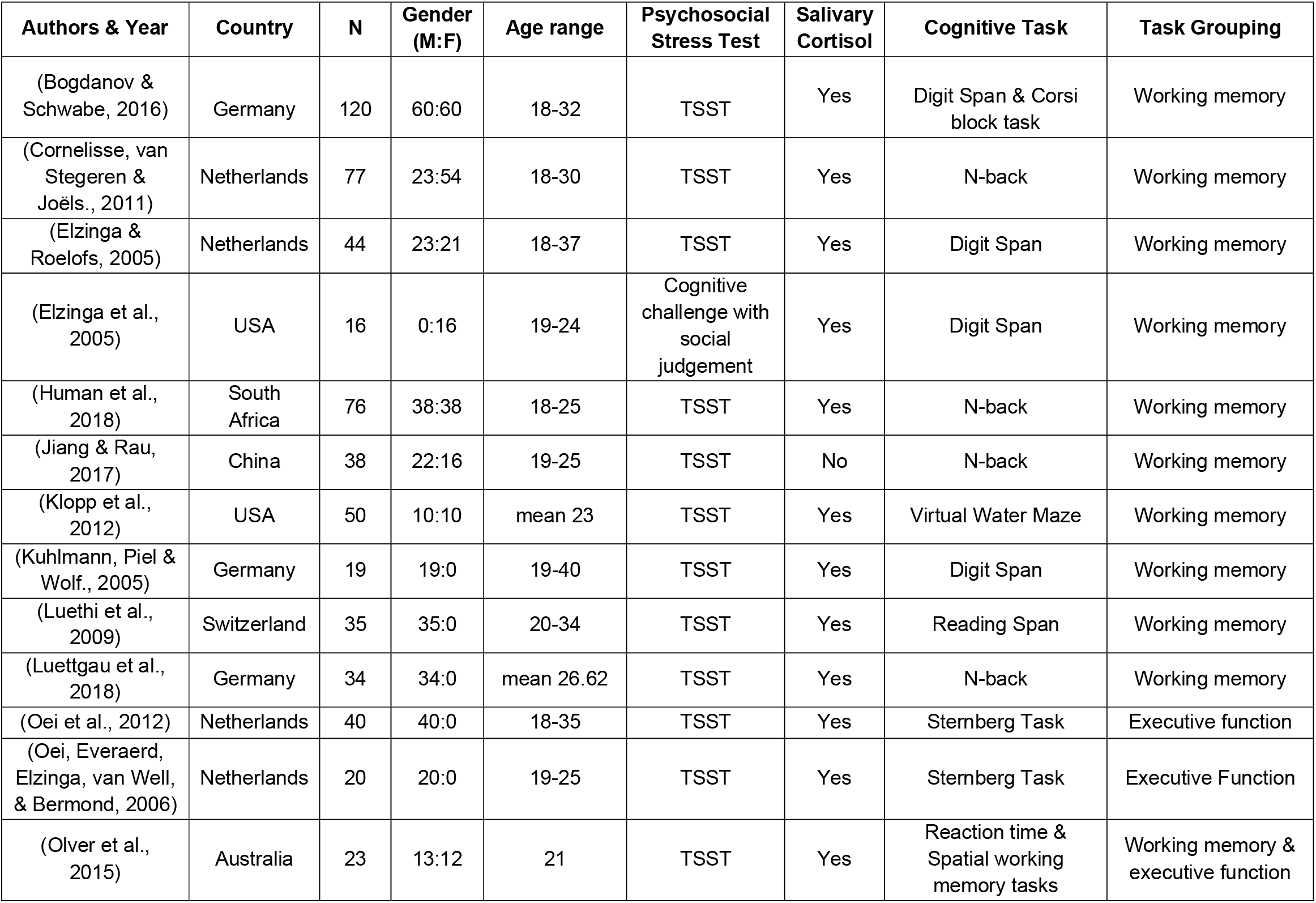
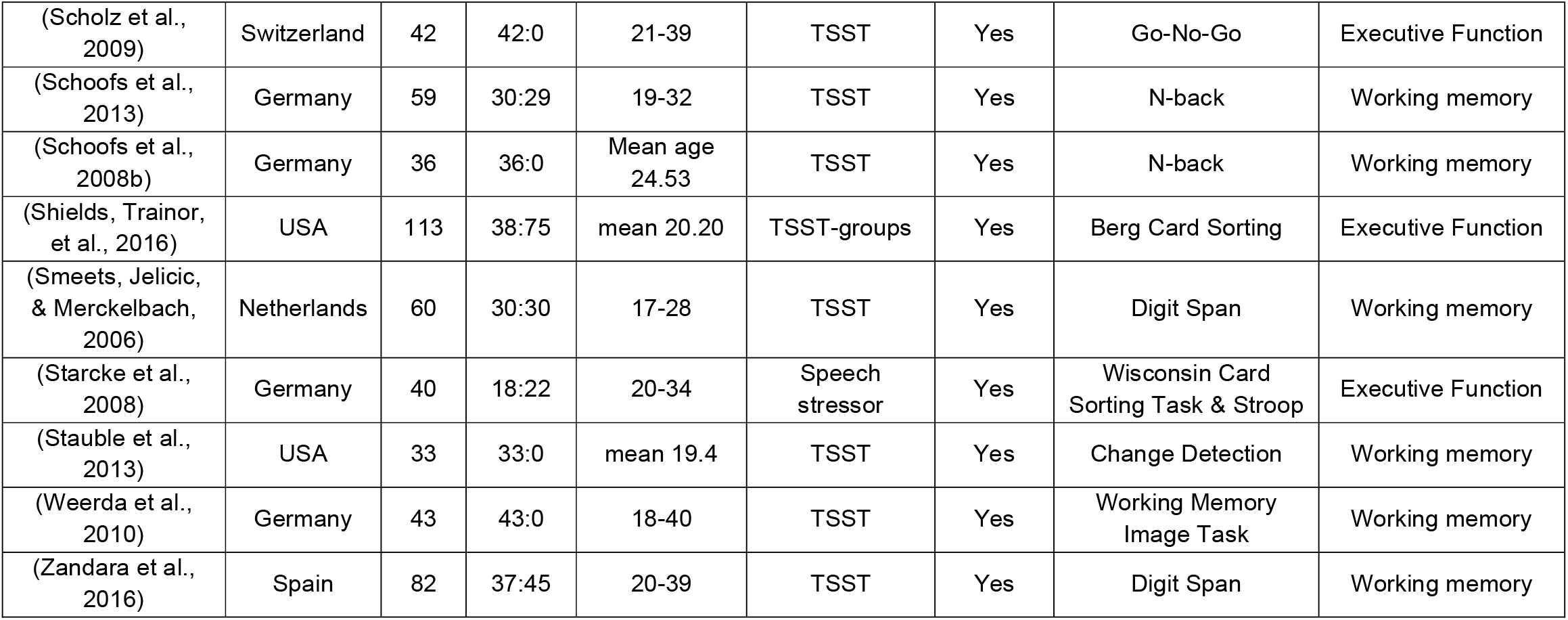
Study characteristics for all included studies using a cognitive functions tasks without testing episodic memory. Task grouping refers to the meta-analysis each study is included in.

The Effects of Psychosocial Stress on Memory and Cognitive Ability: A Meta-Analysis
| Authors & Year | Country | N | Gender (M:F) | Age range | Psychosocial Stress Test | Salivary Cortisol | Cognitive Task | Task Grouping |
| --- | --- | --- | --- | --- | --- | --- | --- | --- |
| (Bogdanov & Schwabe, 2016) | Germany | 120 | 60:60 | 18-32 | TSST | Yes | Digit Span & Corsi block task | Working memory |
| (Cornelisse, van Stegeren & Joëls., 2011) | Netherlands | 77 | 23:54 | 18-30 | TSST | Yes | N-back | Working memory |
| (Elzinga & Roelofs, 2005) | Netherlands | 44 | 23:21 | 18-37 | TSST | Yes | Digit Span | Working memory |
| (Elzinga et al., 2005) | USA | 16 | 0:16 | 19-24 | Cognitive challenge with social judgement | Yes | Digit Span | Working memory |
| (Human et al., 2018) | South Africa | 76 | 38:38 | 18-25 | TSST | Yes | N-back | Working memory |
| (Jiang & Rau, 2017) | China | 38 | 22:16 | 19-25 | TSST | No | N-back | Working memory |
| (Klopp et al., 2012) | USA | 50 | 10:10 | mean 23 | TSST | Yes | Virtual Water Maze | Working memory |
| (Kuhlmann, Piel & Wolf., 2005) | Germany | 19 | 19:0 | 19-40 | TSST | Yes | Digit Span | Working memory |
| (Luethi et al., 2009) | Switzerland | 35 | 35:0 | 20-34 | TSST | Yes | Reading Span | Working memory |
| (Luettgau et al., 2018) | Germany | 34 | 34:0 | mean 26.62 | TSST | Yes | N-back | Working memory |
| (Oei et al., 2012) | Netherlands | 40 | 40:0 | 18-35 | TSST | Yes | Sternberg Task | Executive function |
| (Oei, Everaerd, Elzinga, van Well, & Bermond, 2006) | Netherlands | 20 | 20:0 | 19-25 | TSST | Yes | Sternberg Task | Executive Function |
| (Olver et al., 2015) | Australia | 23 | 13:12 | 21 | TSST | Yes | Reaction time & Spatial working memory tasks | Working memory & executive function |

|  |  |  |  |  |  |  |  |  |
| --- | --- | --- | --- | --- | --- | --- | --- | --- |
| (Scholz et al., 2009) | Switzerland | 42 | 42:0 | 21-39 | TSST | Yes | Go-No-Go | Executive Function |
| (Schoofs et al., 2013) | Germany | 59 | 30:29 | 19-32 | TSST | Yes | N-back | Working memory |
| (Schoofs et al., 2008b) | Germany | 36 | 36:0 | Mean age 24.53 | TSST | Yes | N-back | Working memory |
| (Shields, Trainor, et al., 2016) | USA | 113 | 38:75 | mean 20.20 | TSST-groups | Yes | Berg Card Sorting | Executive Function |
| (Smeets, Jelicic, & Merckelbach, 2006) | Netherlands | 60 | 30:30 | 17-28 | TSST | Yes | Digit Span | Working memory |
| (Starcke et al., 2008) | Germany | 40 | 18:22 | 20-34 | Speech stressor | Yes | Wisconsin Card Sorting Task & Stroop | Executive Function |
| (Stauble et al., 2013) | USA | 33 | 33:0 | mean 19.4 | TSST | Yes | Change Detection | Working memory |
| (Weerda et al., 2010) | Germany | 43 | 43:0 | 18-40 | TSST | Yes | Working Memory Image Task | Working memory |
| (Zandara et al., 2016) | Spain | 82 | 37:45 | 20-39 | TSST | Yes | Digit Span | Working memory |

The samples used in the included papers were primarily European. Of these European papers, 21 collected their samples in Germany, 8 in The Netherlands, 3 in Spain and 4 in Switzerland. Several studies also used samples from North America, with 9 from the USA and 2 from Canada. The final four studies were carried out in Japan, China, South Africa and Australia. The majority of studies included in the review were published within the last 15 years.

### 3.2 Stress & Memory Tasks

Psychosocial stress was induced using the Trier Social Stress Task (TSST) (Kirschbaum, Pirke, & Hellhammer, 1993) in 43 of the papers included in this review. One further study used an adapted TSST for group’s stressor. Two studies using a version of the Montreal Imaging Stress Task (MIST), four used a public speaking stressor and the final study used a complex cognitive challenge with social judgement. Physiological responses to stressors were measured using saliva samples to quantify elevations in cortisol in 49 of the 51 included papers.

Memory was assessed using free recall in 21 papers, and recognition in 11 papers. Three papers measured memory using both tasks. Separate effect sizes were calculated for each task and both were included. Similarly, there was variability in the type of task used to measure other cognitive functions. Some studies used several tasks to explore multiple cognitive functions. Working memory was assessed using the n-back task in seven papers, reading and digit span tasks in eight papers and visuo-spatial tasks in four further papers. Attention was assessed in three papers, and cognitive flexibility was explored in four other papers, all using a variety of different tasks. As these tasks are independent, effect sizes for each task were calculated and included in the relevant analysis.

### 3.3 Risk of bias

Two elements within the CASP were excluded: (1) all participants had to complete the full study for their data to be included and (2) given we explicitly considered time between sessions in this review, we chose not to assess the quality of studies based on follow up criteria. Sample size calculations were reported in only a few studies, suggesting there may be potential for underpowered studies skewing results (albeit with both false positives and false negatives). We therefore include risk-of-bias analyses to mitigate against such issues. The results of the quality assessment using the CASP suggest that the studies included in this review were generally of a high standard with 36 papers being rated as good and 15 rated as fair.

One difficulty with comparing the studies included in this review has been the relatively low number of studies included in specific analyses. There were too few studies examining post-encoding stress with long retention intervals (category iv; 3 studies) and stress during encoding (category vi; 2 studies); any meta-analysis performed on this data could be misleading (Valentine et al., 2010). Instead, we offer a narrative discussion of these studies. Exact numbers of studies included in each analysis conducted can be found in Table 3.

**Table 3.**
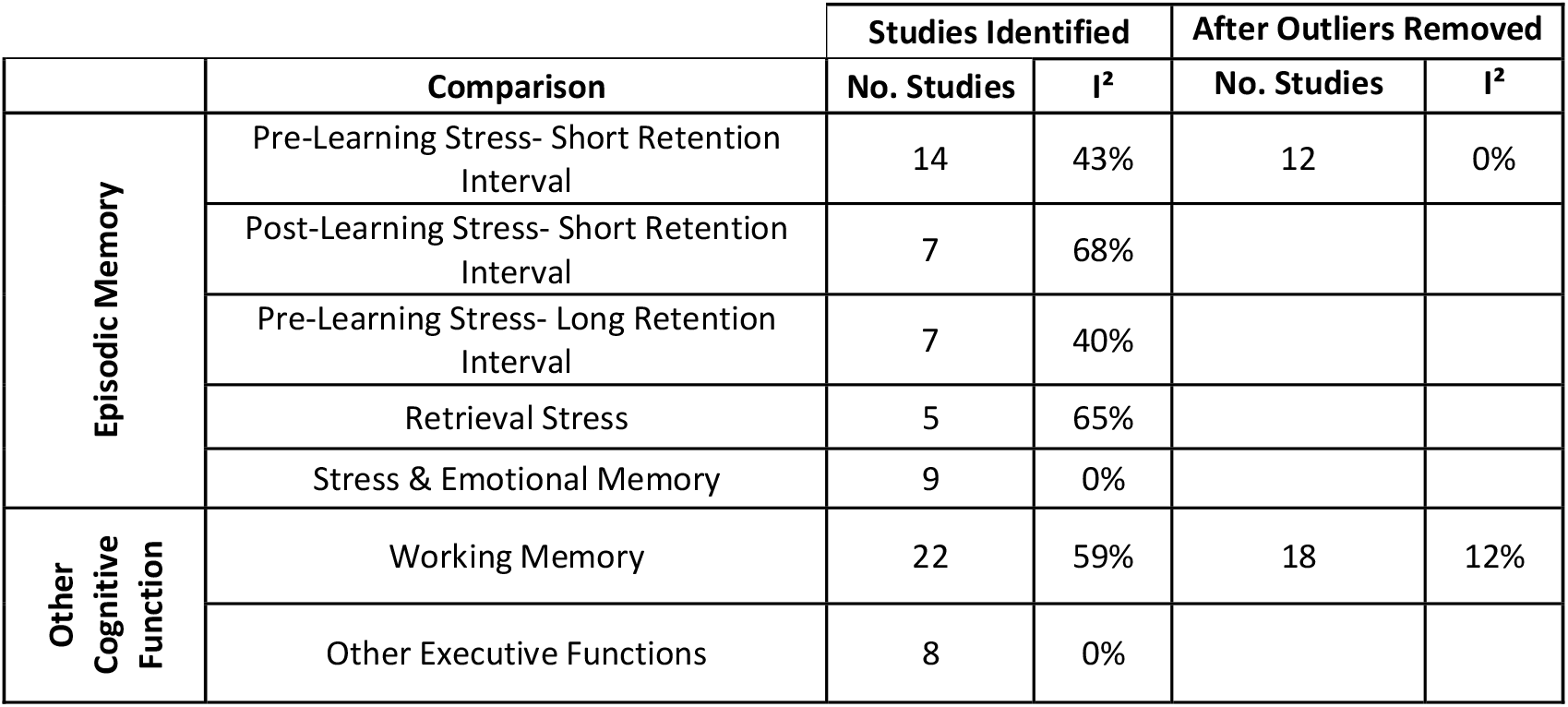
Heterogeneity of meta-analyses using I^2^ statistic to describe the percentage of variation across studies that is due to heterogeneity rather than chance. Values are given for the number of studies initially identified for each grouping and for final totals after outliers identified and removed based on funnel plots and sensitivity analysis.

|  | Comparison | Studies Identified |  | After Outliers Removed |  |
| --- | --- | --- | --- | --- | --- |
|  |  | No. Studies | I<sup>2</sup> | No. Studies | I<sup>2</sup> |
| Episodic Memory | Pre-Learning Stress- Short Retention Interval | 14 | 43% | 12 | 0% |
|  | Post-Learning Stress- Short Retention Interval | 7 | 68% |  |  |
|  | Pre-Learning Stress- Long Retention Interval | 7 | 40% |  |  |
|  | Retrieval Stress | 5 | 65% |  |  |
|  | Stress & Emotional Memory | 9 | 0% |  |  |
| Other Cognitive Function | Working Memory | 22 | 59% | 18 | 12% |
|  | Other Executive Functions | 8 | 0% |  |  |

In order to assess risk of bias using funnel plots, comparisons must include 10 //studies (Sterne et al., 2011). Three of the seven analyses included more than 10 studies. Two analyses removed multiple studies on this basis: two studies were removed from pre-learning stress-short retention interval analysis (Smeets et al., 2006, 2012) and four studies were removed from the working memory analysis (Cornelisse et al., 2011; Jiang & Rau, 2017; Luethi et al., 2009; Stauble et al., 2013). Figure 2 shows funnel plots before and after outliers have been removed from two of the relevant analyses.

**Figure 2.**
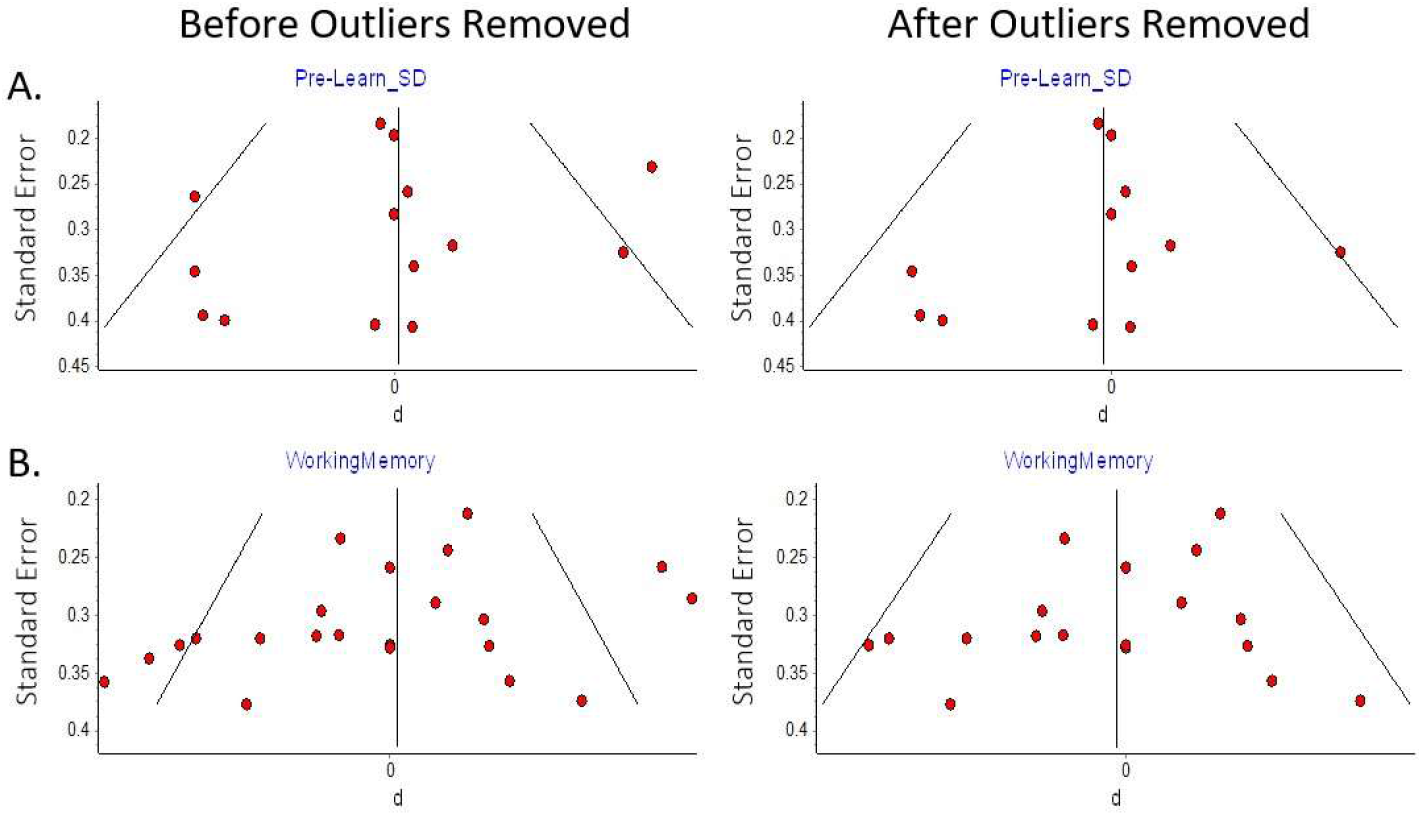
Risk of Bias. Funnel plots with and without outliers removed based on sensitivity analysis.

**Figure 3.**
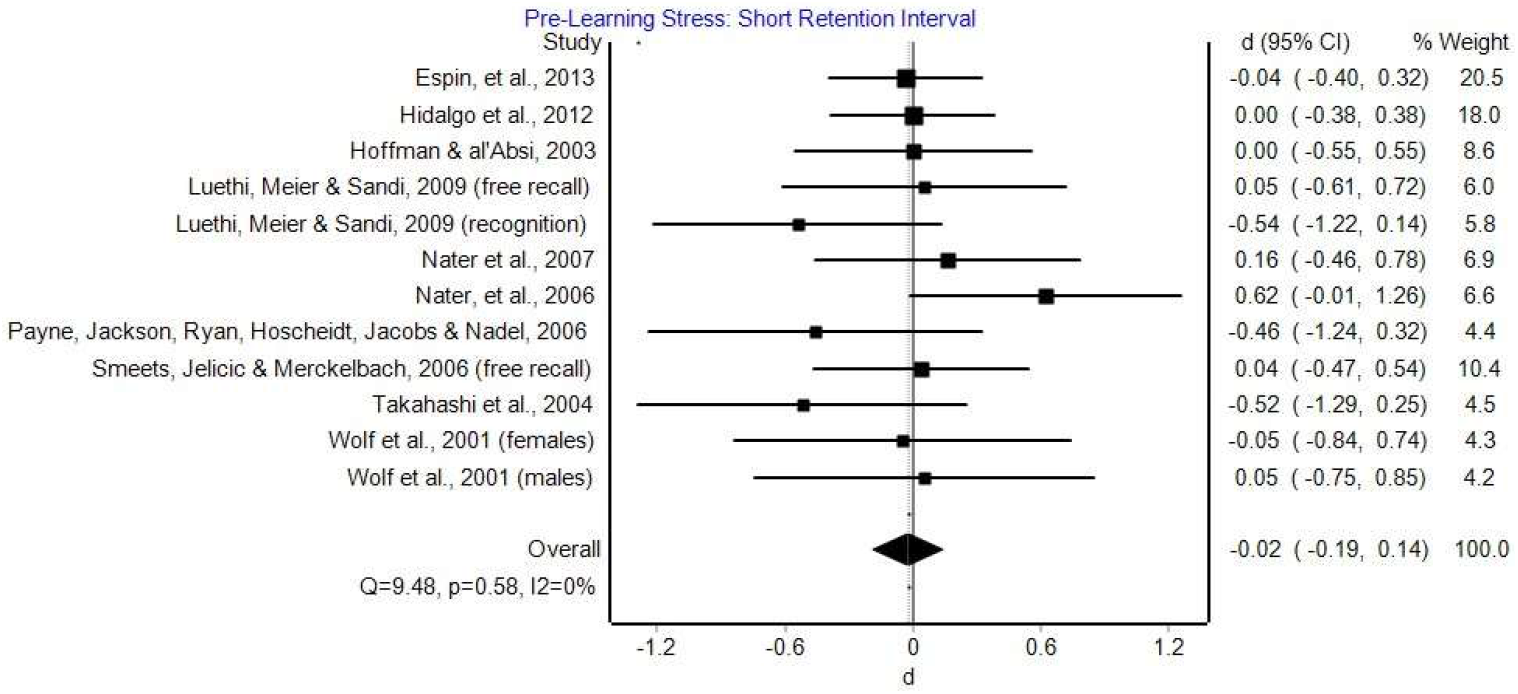
Pre-learning psychosocial stress with a short retention interval. Effect sizes and 95% CI’s for the effects of pre-learning stress on memory.

**Figure 4.**
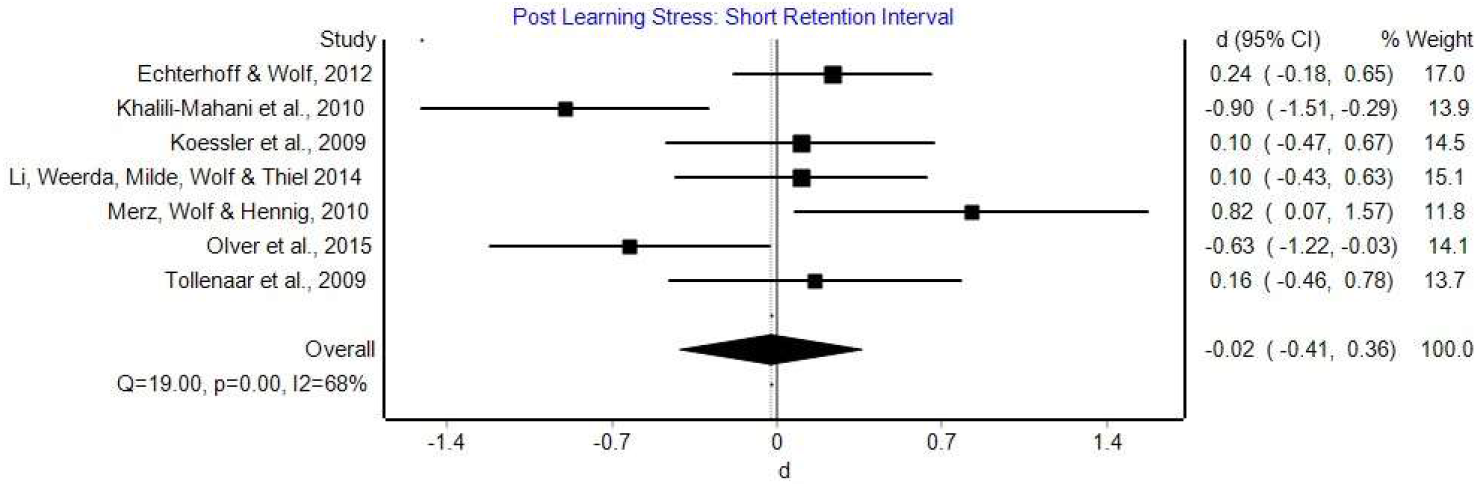
Post learning psychosocial stress with a short retention interval. Effect sizes and 95% CI’s for the effects of post-learning psychosocial stress on memory.

**Figure 5.**
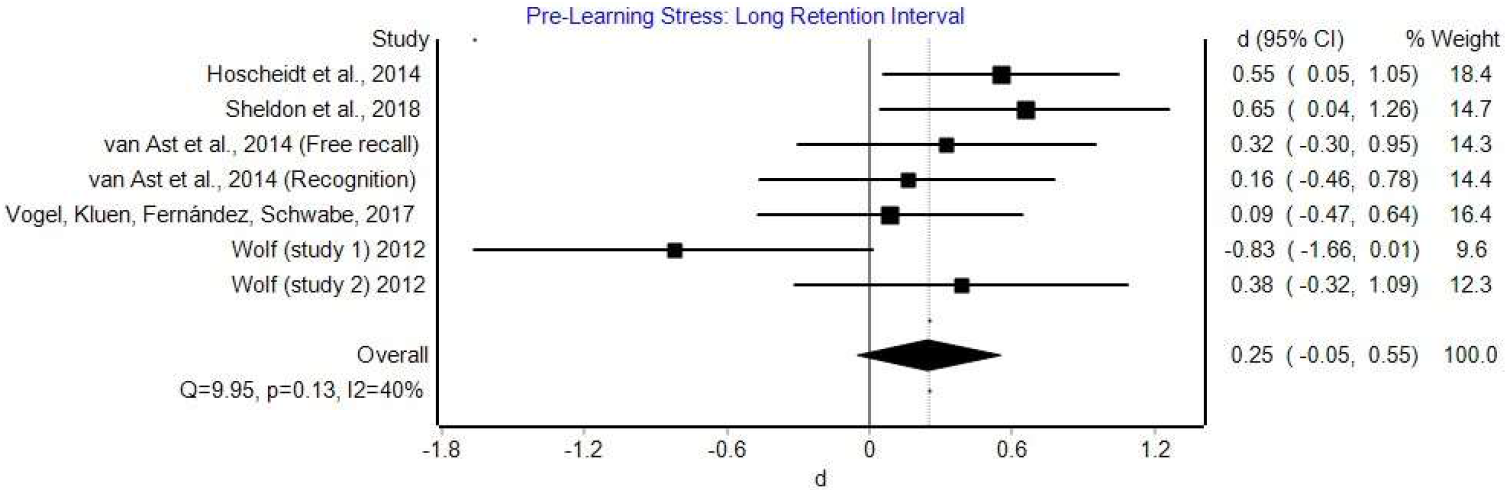
Pre-learning psychosocial stress with a long retention interval. Effect sizes and 95% CI’s for the effects of pre-learning stress on memory with at least a one-day delay period between stress and memory retrieval.

**Figure 6.**
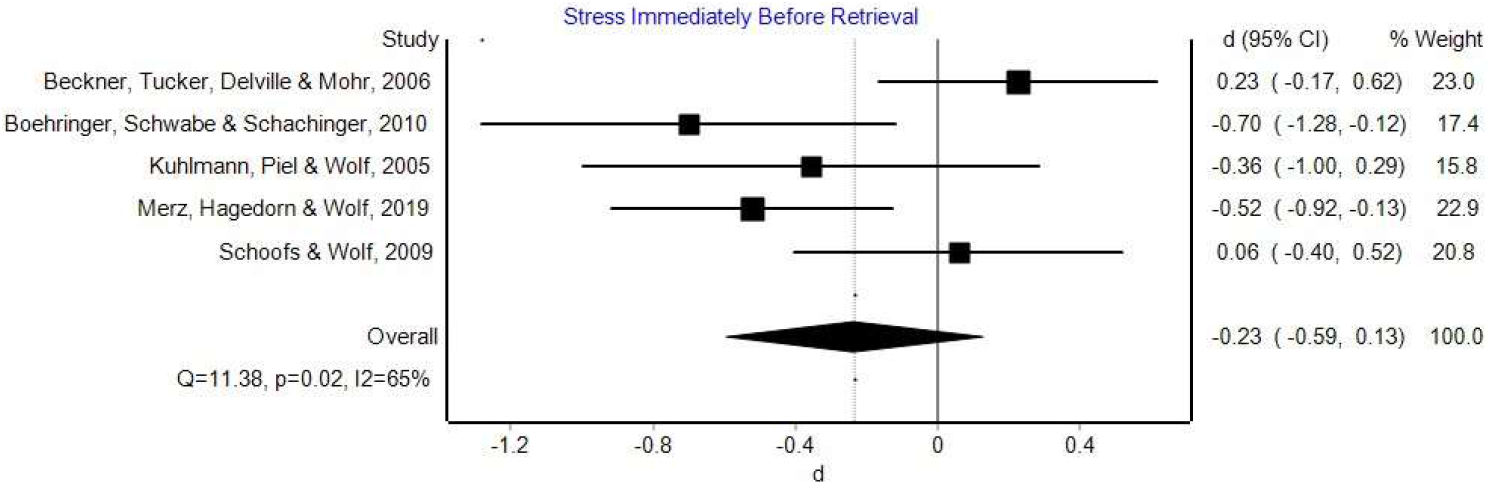
Psychosocial stress immediately before retrieval. Effect sizes and 95% CI’s for the effects of stress on retrieval.

**Figure 7.**
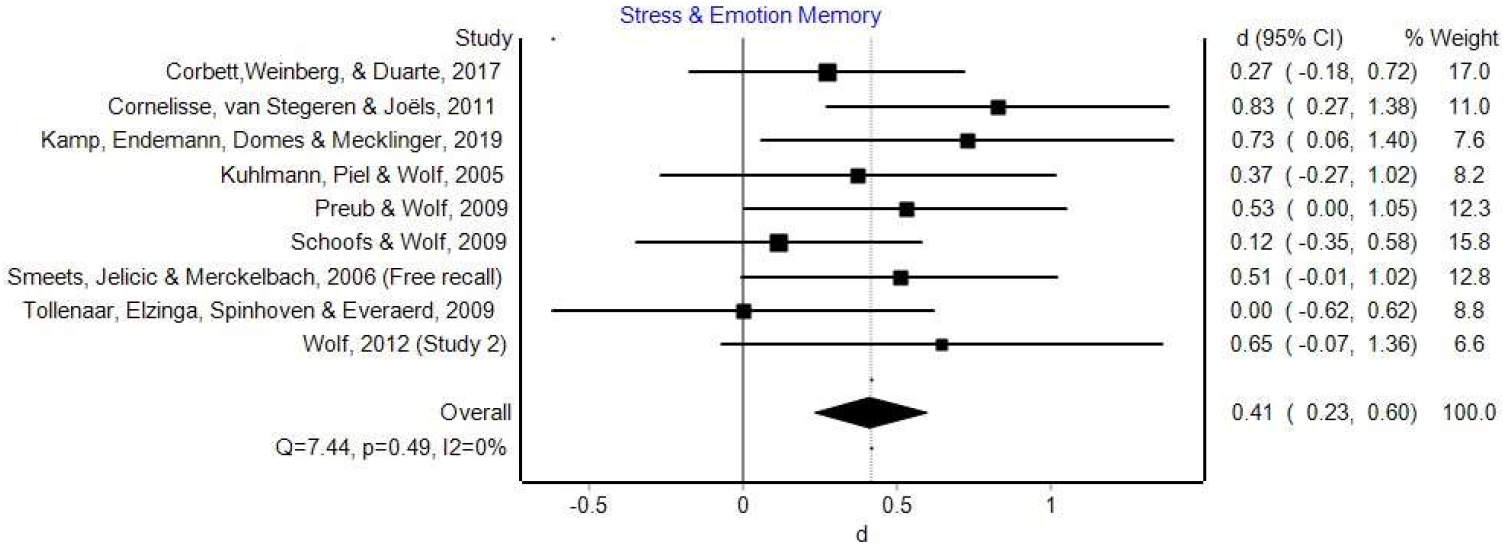
Interaction between psychosocial stress and stimuli type (emotional vs neutral). Effect sizes and 95% CI’s for the effects of stress on emotional memory.

**Figure 8.**
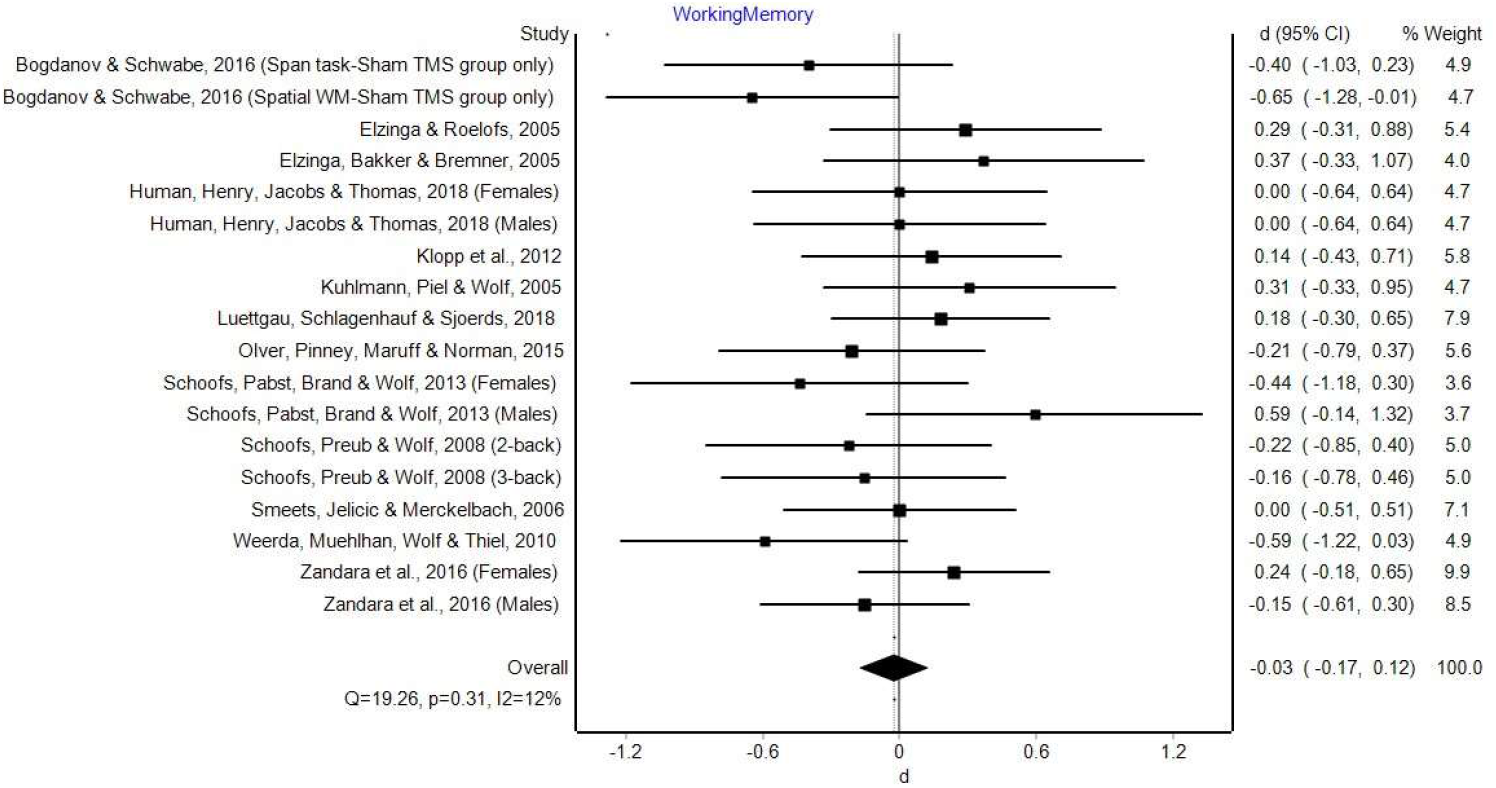
Psychosocial stress and working memory. Effect sizes and 95% CI’s for the effects of stress on working memory.

**Figure 9.**
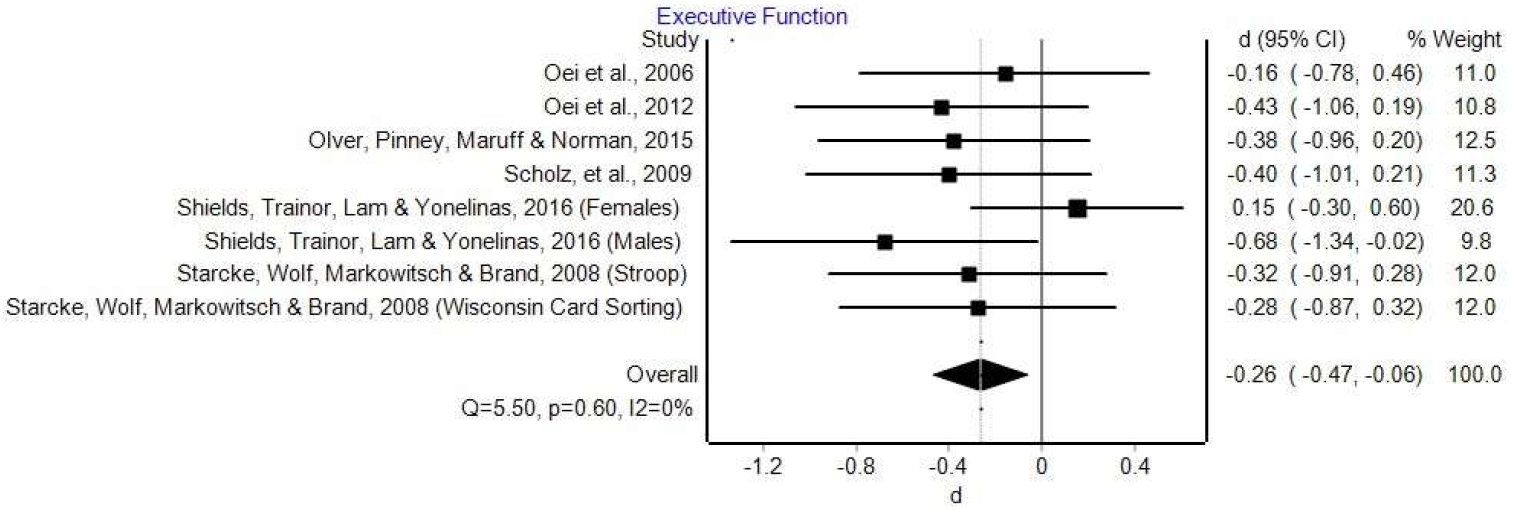
Psychosocial stress and executive function. Effect sizes and 95% CI’s for the effects of stress on executive function.

The results of the heterogeneity analysis are shown below (Table 3). Before splitting studies by stress and memory phase, all studies assessing episodic memory showed moderately high heterogeneity (I^2^ = 56%), suggesting the need to split studies into more relevant comparisons. Based on a cut off points for I^2^ values of <40% (Higgins & Thompson, 2002), the majority of meta-analyses we conducted scored low for heterogeneity. The two analyses with higher scores of heterogeneity were also two of those which included the smallest number of studies, and therefore the I^2^ score could be inflated by lower study numbers (Higgins & Thompson, 2002).

### 3.4 Psychosocial Stress and Episodic Memory

We first examined the main effect of psychosocial stress on different stages of memory, before considering effects on emotional and neutral stimuli.

#### 3.4.1 Effects of Timing of Stress Induction on Memory

##### 3.4.1.1 Pre-learning stress with a short retention interval

Twelve studies were included in this analysis. We found mixed effects of psychosocial stress on encoding in this design, with eight studies showing no effect, three medium negative effects and one medium positive effect. Overall no effect was found (Cohen’s d = -0.02, 95% CI’s = -0.19-0.41).

##### 3.4.1.2 Post-Learning stress with short retention interval

Seven studies were included in this analysis. Four found no effect of post-learning psychosocial stress on memory. One study found a positive small effect and another showed a large positive effect. The remaining two studies found negative effects (one medium, the other large). Overall there was no significant effect of post learning stress on retention after a short interval (Cohen’s d = -0.02, 95% CI’s= -0.41-0.36).

##### 3.4.1.3 Pre-Learning stress with long retention interval

Seven studies were included in this analysis. One study found a large negative effect, while four studies showed small and medium positive effects. The remaining two studies showed no significant effect. Overall, there was a small, non-significant, effect of pre-learning psychosocial stress on memory when using a long retention interval (Cohen’s d = 0.25, 95% CI’s = -0.05-0.55).

##### 3.4.1.4 Post-Learning stress with long retention interval

Three studies were included in this analysis. Due to low study numbers and high variability, any meta-analysis of this data is unlikely to provide meaningful results. Two studies suggest positive effects of post-learning psychosocial stress on subsequent memory retrieval and are included in later meta-analyses (Corbett et al., 2017; Preuß & Wolf, 2009). The remaining study showed a medium positive effect of stress on memory d = 0.59 (Beckner et al., 2006). Together, these findings suggest that post-encoding psychosocial stress may enhance recognition memory.

##### 3.4.1.5 Stress immediately before retrieval

Five studies examined the effects of pre-retrieval stressors after a long *retention interval*. Three studies found that stress immediately before retrieval significantly impaired memory, with small and medium effect sizes, one study showed a small positive effect, while the remaining study suggested no effect of stress on memory. Overall, there was a small, non-significant small effect of psychosocial stress immediately before retrieval on memory (Cohen’s d = -0.23, 95% CI’s = -0.59 --0.13).

##### 3.4.1.6 Stress during encoding

Two studies induced psychosocial stress during encoding but could not be compared using a meta-analysis. Both of these studies suggested that stress enhanced memory for the central aspects of the information they encoded relative to control groups with a small, d = 0.25 (Wiemers et al., 2013), and a medium effect size, d = 0.74 (Herten et al., 2017).

#### 3.4.2 Emotional Memory

Nine studies examined the effects of psychosocial stress on memory for emotional and neutral stimuli. Effect sizes were calculated using the interaction between condition (stress vs. control) and stimuli type (emotional/ neutral). Comparing studies in this way can only reveal if there is an effect of stress on emotional compared to neutral stimuli, but not whether stress particularly impacts memory for stimuli with a specific valence, or the direction of that effect. Two studies showed no effect, two showed a small effect, four a medium effect and one a large effect for an interaction between psychosocial stress and stimuli type (emotional or neutral) with, overall, a significant small effect (Cohen’s d = 0.41, 95% CI’s = 0.23-0.6).

The effect of psychosocial stress on memory for the specific valence of emotional stimuli was inconsistent. Three studies showed no significant interaction between stimuli valence and stress condition, suggesting psychosocial stress has similar effects on material of all valences. One study reported that stress enhanced memory for both positive and negative emotional stimuli relative to neutral, but another found the opposite effect: both positive and negative emotional stimuli were less well remembered than neutral words following stress. Similarly, one study found that memory for positive emotional stimuli was enhanced by social stress, two found that memory for negative emotional stimuli was enhanced by stress. One further study also found that memory for negative emotional words was specifically impaired following stress. This inconsistency in findings gives no clear indication as to specifically how emotional memory is influenced by psychosocial stress.

### 3.5 Psychosocial stress and other cognitive functions

#### 3.5.1 Working memory

Eighteen studies assessed working memory using at least one of the three different task types; digit/reading span, spatial and n-back tasks. When considering working memory overall, no significant effect of psychosocial stress on working memory was found (Cohen’s d = -0.03, 95% CI’s = -0.17-0.12).

#### 3.5.2 Executive function

Eight studies assessed the influence of psychosocial stress on different executive function tasks. Five studies showed a small negative effect, one showed a negative medium effect and a further one showed a very small negative effect. One final study showed a very small positive effect. Overall there was a significant negative small effect of psychosocial stress on executive function (Cohen’s d = -0.26. 95% CI’s = -0.47--0.06).

## 4. Discussion

The aims of this systematic review and meta-analysis were to explore the effects of psychosocial stress on (1) different stages of memory, (2) memory for emotional stimuli, and (3) other cognitive functions that influence long-term memory. This meta-analysis suggests little to no effect of psychosocial stress on episodic memory regardless of time of stressor. There is some indication that stimuli valence may influence vulnerability to stress. Additionally, although stress has limited effects on working memory, other executive functions were found to be impaired.

### 4.1 Timing of stress

Psychosocial stress had minimal effect on each of the memory stages. This was the case whether or not studies included a delayed period between learning and retrieval. Despite variability in the literature, as a whole, studies overall showed no significant effects of psychosocial stress on memory for any of these comparisons. The inconsistency in findings may reflect the heterogeneity of study designs and potentially unaccounted for differences in samples. These, and possible as yet undefined other factors (including inter-individual variability), may influence the variable findings for studies assessing the effects of psychosocial stress on memory.

Our findings differ from previous reviews exploring the effects of all forms of stress on memory, as opposed to psychosocial stress alone (Shields et al., 2017). This discrepancy may suggest differences in how particular forms of stress impact upon memory. As noted in previous reviews, psychosocial and physical forms of stress are associated with increased activity in different regions of the brain (alongside some overlap), with varying degrees of involvement of key memory regions, such as the hippocampus, amygdala and prefrontal cortices (Dedovic et al., 2009). The extent of activation of these regions by specific forms of stress could linearly relate to stress-related memory alterations, which could be tested in future studies.

### 4.2 Effect of psychosocial stress on memory for emotional stimuli

Relatively few studies have examined how psychosocial stress impacts upon memory for emotional compared to neutral stimuli. These studies found, overall, a small significant effect. Importantly, whilst there was an overall effect of stress on stimuli type (emotional compared to neutral), the direction of this effect, and the impact on stimuli with a specific valence, was ambiguous. This inconsistency is important to consider. If we are to understand the mechanisms underlying why psychosocial stress might particularly impact memory for emotional stimuli, and create suitable models for this, then this needs to be done on a foundation of consistent results. It may be factors like specific memory tasks influence these outcomes but, in that case, it needs careful systematic exploration in well-designed studies. In turn this could help unravel the mystery of why some studies show clear effects of stress on ‘memory’ and others do not. It is therefore likely an oversimplification to state that stress impacts memory for emotional stimuli through its effects on specific neurobiological routes, such as through additional recruitment of the amygdala (Cahill et al., 2003; McCullough & Yonelinas, 2013; McGaugh, 2004; Yonelinas & Ritchey, 2015). The studies which did report a memory advantage for emotional stimuli all administered stress prior to encoding. Additional amygdala activation exclusively during consolidation or prior to retrieval is therefore unlikely to be enough to facilitate this memory advantage.

### 4.3 Stress Responsivity

Within this review, very few studies explored inter-individual differences in cortisol responsivity. However there is evidence to suggest that these differences may be of particular importance (Oei et al., 2006; Takahashi et al., 2004; Wolf et al., 2001). All three of these studies found a significant, large effect of psychosocial stress negatively impacting memory when carried out immediately before retrieval in those with higher cortisol reactivity. There were too few studies to confirm this in a meta-analysis but the consistency in findings is of note. It could be that inter-individual differences play a greater role in determining the effects of stress on memory than simply psychosocial stress alone.

### 4.4 Effects of psychosocial stress on other cognitive functions

We also asked why memory might be affected by psychosocial stress; specifically, whether other cognitive functions, such as working memory or executive function, mediate the relationship. In line with our other analyses we found little evidence for a consistent effect of stress on working memory. This finding differs from earlier studies suggesting that mainly physical stress (but also to a lesser extent psychosocial stress) impairs working memory (Schoofs, Wolf & Smeets, 2009; Shields, Sazma & Yonelinas, 2016).

For executive function, there was a small, significant, negative effect of psychosocial stress. Stress was found to impair tasks like attention or cognitive flexibility, such as the Wisconsin Card Sorting Test (Sänger et al., 2014; Shields, Sazma & Yonelinas 2016). It could be argued that effects on executive function dictate whether studies find an effect of psychosocial stress on memory: tasks that relay on strategic retrieval or encoding process (such as tests of recall or complex stimuli) may be more sensitive. The impact on executive function and indirect effects on memory could be carefully examined in future studies to determine exactly what practical implication these effects have.

### 4.5 Limitations

Although attempts were made to compare the effects of psychosocial stress as fairly as possible and to minimise any possible bias, there was still some issues of heterogeneity in analyses with lower numbers of studies. In these cases, findings should be interpreted with caution (e.g. studies examining the effects of stress immediately before retrieval). In some cases, the number of studies were too low to perform a meta-analysis. For example, we were unable to compare the effects of post-encoding stress with long-retention intervals on memory. Despite these concerns, we identified 36 studies assessing the effects of psychosocial stress on long-term episodic memory, and from these a consistent picture emerges of only limited effects, regardless of the timing at which stress occurs. In future, we would strongly encourage multi-experiment designs, that attempt to pinpoint exactly which aspects of memory are affected by psychosocial stress. It may be particularly important to define the stress reactivity of samples, to fully understand the mechanisms underpinning this relationship.

### 4.6 Conclusion

In this review we examined the specific effects of psychosocial stress on memory. Unlike previous studies, we did not conflate physical and psychosocial stress. As a result we found much more limited effects on memory than has previously been reported (e.g. Shields et al., 2017). This finding was not affected by the timing at which stress was applied (e.g. before encoding or retrieval). Executive functions, but not working memory, were impaired following psychosocial stress. This could potentially mediate the proposed relationships between psychosocial stress and memory, but it still needs to be determined whether tasks that rely on controlled processes are more sensitive to psychosocial stress. Psychosocial stress did have differential effects on memory for neutral and emotional stimuli, but the direction of this effect was inconsistent, and was mainly seen when stress was carried out prior to encoding. This field is complex, with many more moving parts involved in studies than may be appreciated. We cannot assume that what seems to be true for physical stressors or pharmacological administrations of stress-related hormones applies to psychosocial stress. This is critical when attempting to infer the relevance of these studies to workplace or other real-world situations, which predominantly involve responses to psychosocial stressors in the environment.

## Data Availability

Data is available upon reasonable request

## 5. Acknowledgements

